# Assessment of WBCT Use in Trauma Patients: A National audit for Radiology Resource Management and Limiting Radiation Exposure

**DOI:** 10.1101/2025.04.09.25325527

**Authors:** Latifah Al-Kandari, Michael Masoomi, Abdelhamid El Gargani, Mahdy Abass Hamza, Rami Mohamed Agha

**Affiliations:** Department of Radiology, Adan Hospital, Al-Mosely Street, Hadiya, 46969, Ministry of Health, KW; Department on Nuclear Medicine and Molecular Imaging, ADAN Hospital, Hadiya, 46969, Ministry of Health, KW

## Abstract

**Introduction:** Whole-body computed tomography (WBCT) is often indiscriminately used in trauma cases. This study aimed to assess the effective use of WBCT in trauma patients across emergency departments in State of Kuwait, evaluate the true incidence of critical injuries, and minimize unnecessary radiation exposure.

**Methods:** This multicenter, retrospective study across seven radiology departments included 1,367 trauma patients who underwent WBCT between 2022 and 2023, according to the American College of Radiology guidelines. Data on age, sex, injury mechanism, clinical indications, dose-length product, and WBCT findings were collected and analyzed using IBM SPSS version 25.

**Results:** Of 1,367 referrals, 578 (42.3%) had no significant findings, while 789 (57.7%) showed positive trauma-related results. Among the positive findings, 530 patients (38.8%) had major injuries, including solid organ and vertebral column injuries. The most common causes of WBCT referrals were road traffic accidents (RTAs) (911 patients, 66.6%), falls from height (FFH) (182 patients, 13%), falls of heavy objects (112 patients, 8%), head trauma (82 patients, 6%), buggy accidents (28 patients, 2%) and others. Negative WBCT findings had a mean effective dose (ED) of 19.98 ± 10.26 mSv.

**Conclusions:** Positive WBCT findings were seen in 57.7% of cases, but the 42.3% negative findings rate, was significantly higher than the 20.4% average reported in similar centers in Europe such as UK and Germany, which highlights the need to optimize WBCT usage in emergency department to reduce unnecessary scans and minimize radiation exposure.

## 1. Introduction

Major trauma is the sixth leading cause of death worldwide and the leading cause of death and disability in the population aged 5 to 45 years [1]. The assessment is typically guided by strict protocols to quickly identify life-threatening conditions before conducting a comprehensive evaluation of other injuries. These protocols often prioritize the use of plain film radiography, ultrasonography and laboratory testing over comprehensive history and physical examination. Common initial imaging strategies include chest and pelvic radiographs and Focused Assessment with Sonography for Trauma (E-FAST), which are cost-effective and easy to perform but offer limited diagnostic accuracy. Several studies have shown that a thorough clinical examination, including the use of FAST, chest X-rays, and pelvic X-rays, can effectively exclude injuries, potentially avoiding unnecessary WBCT scans [2-3]. In contrast, whole-body computed tomography (WBCT) is often used as the primary assessment tool in trauma centers due to its high sensitivity and speed in detecting traumatic injuries [4].

In severely injured patients, WBCT provides accurate details about the extent of injuries and is also used to identify occult injuries in high-energy trauma patients who may not show obvious signs of injury [2, 5-8]. The rationale for routine WBCT use is supported by studies showing a high risk of missing significant injuries in trauma patients without it. However, the routine use of WBCT in major trauma patients, even when no injury is clinically suspected, remains controversial, particularly from a radiation protection standpoint [3].

Studies have indicated that 1–3% of cancers globally are attributed to medical sources of radiation [9-11]. On average, WBCT exposes each patient to more than 20 mSv of effective radiation, which increases the risk of cancer mortality [3]. For a 35-year-old male, the risk of cancer mortality is estimated at 1 in 900 with a radiation dose of 24 mSv, while for an average 45-year-old adult, the risk is 1 in 1,250 with a radiation dose ranging from 10–20 mSv. [9,12-14].

While WBCT use in State of Kuwaitis aligned with the country’s broader healthcare objectives under State of Kuwait Vision 2035, which aims to modernize the healthcare system and improve patient outcomes, optimizing WBCT use is critical to achieving these goals without overburdening healthcare resources or exposing patients to unnecessary radiation risks. WBCT is widely available in government hospitals under the Ministry of Health (MOH). Its use is common in trauma cases, particularly for road traffic accidents (RTAs), which are a major public health issue in State of Kuwait[15].

Recently, there has been a shift from indiscriminate WBCT referrals for trauma patients, particularly those with high-energy trauma, towards a more risk-benefit-oriented approach. This approach incorporates clinical prediction rules to safely exclude WBCT in certain cases [16]. The aim of this study was to estimate the true incidence of clinical injuries referred to emergency departments at radiology centers in Kuwait. We hypothesised that this estimation could lead to more effective utilization of radiology resources, which are strained by the high volume of referral patients and could contribute to the optimisation of radiology healthcare services.

## 2. Methods

This multicentric retrospective study included all seven radiology departments in Kuwait, examining 1367 trauma patients from 2022-2023 who referred from emergency departments. All trauma patients were initially managed and proceed to WBCT in accordance with the American College of Radiology guidelines for Major Blunt Trauma [17].

### 2.1 Inclusion and Exclusion Criteria

All patients with major traumatic event, hemodynamic stability and altered consciousness who were admitted to the emergency department were included in this study and underwent WBCT. Any other causes were excluded from this assessment.

### 2.2 Data Collection

The proposed study received approval from the Ethics Committee of the relevant institution (MOH -KEC 1298). Data were collected from trauma patients admitted to the emergency departments of collaborative radiology centers. Consent was obtained as part of the standard clinical study. Three of the seven departments were equipped with GE CT-128, GE CT-256, and Siemens CT-128 scanners, while the remaining had Siemens CT-256 scanners. Patients received intravenous (IV) contrast-medium for the chest and abdomen. Post-imaging, all patient data were archived using PACS systems, with four departments utilizing Centricity, two using Agfa, and one using Siemens.

Age, sex, mechanism of injury, clinical requests, clinical findings, and WBCT findings in the cervical spine, thoracolumbar spine, chest, abdomen, pelvis, and appendicular skeleton were recorded based on institutional reporting. Radiological results were initially classified into negative and positive findings. Positive findings were then categorized as major or minor injuries based on impact of energy, physiological disturbances, potential for life-threatening conditions and anatomical regions involved. The specification of WBCT scan acquisition parameters, dose-length product (DLP) values from the scanner-generated dose reports and a conversion factor that is, the region-specific normalized effective dose per DLP (mSv × mGy−1 × cm−1) [18] were used to calculate the effective dose (ED). The ED was estimated as the product of the DLP and the corresponding conversion factor (k): ED (mSv) ≈ k × DLP. For the whole-body scan, we used a k value (k=0.0093 mSv × mGy−1 × cm−1) [19-20]. CT Automatic exposure control (AEC) that modulates radiation exposure automatically and is widely used for optimization of radiation dose in CT were used by the departments [21-22]. The Sante DICOM viewer was utilized to retrieve all the relevant parameters.

### 2.3 Statistical Analysis

A power calculation with a 10% margin of error and 95% confidence level indicated a minimum of 96 datasets per center. However, 200 WBCT trauma patient datasets were collected from each center for a more robust analysis. Pre-test clinical requests, including clinical signs, symptoms, injury mechanisms, and clinical queries, were compared with WBCT findings. Data analysis was conducted using IBM SPSS version 25.0. The Kolmogorov-Smirnov test assessed normality. Statistical significance was set at the 5% level. Chi-square, Student t-test, and Mann Whitney tests were used to compare categorical and quantitative datasets.

## 3. Results

A total number of 1,367 cases were included and assessed in this study. The mean age of patients referred to all cohort departments was 33.54± 16.09 years, where 1,074 (78.6%) were males and 293 (21.4%) were females, and male to female ratio was 3.7:1. Out of 1,367 patients scan, 578 (42.3%) were reported as normal (negative finding), whereas 789 patients (57.7%) had positive trauma related radiological finding of which 530 patients were found to have a major injury (38.8%) including solid organ and vertebral column injuries (Table 1). According to CT indication, 911 patients had been referred due to RTA (66.6%), 182 patients due to FFH (13%), 112 patients due to a fall of a heavy object (8%), 82 patients due to head trauma (6%), while 28 patients had injuries due to buggy accident (2%), and 24 patients had an injury due to assault (1.8%). The RTA, with a 66.6 % score was the most common indication for WBCT referral (Table 2), (Figure 1). Anatomical body regions that were affected according to the WBCT scan results were Head (48.7%), chest (31.1%), abdominal (10.2%), pelvis (7.6%), others (2.4%) (Figure 2).

**Table 1.** Demographic characteristics and classification findings.

|  |  | Total cases<br>(n=1367) | Negative scan<br>(n=578) | Positive scan<br>(n=789) | p-value<sup>‡</sup> |
| --- | --- | --- | --- | --- | --- |
| Gender (%) | Male | 1074 (78.6%) | 427 (73.9%) | 647 (82.0%) | <0.001<sup>‡</sup> |
|  | Female | 293 (21.4%) | 151 (26.1%) | 142 (18.0%) |  |
| Age | Mean<br>±SD | 33.54± 16.09 | 32.03 ±15.97 | 34.63 ±16.09 | <0.001<sup>‡</sup> |
| Finding Classification: |  |  |  |  |  |
|  | Major |  |  | 530 (38.8%) |  |
|  | Minor |  |  | 259 (18.9%) |  |

<sup>‡</sup> Mann-Whitney U test, <sup>‡</sup> Chi-Square Test

**Table 2.** Mechanism of injury and WBCT indications-rate of positive and negative scans.

| Mechanism of Injury | Total Cases<br>(n=1367) | Negative Scan<br>(n=578) | Positive Scan<br>(n=789) | *p*-value<sup>‡</sup> |
| --- | --- | --- | --- | --- |
| RTA | 911 (66.6%) | 405 (70.1%) | 506 (64.1%) | 0.376 |
| FFH | 182 (13.6%) | 42 (6.7%) | 140 (17.7%) | <0.001 |
| Fall (fall of heavy object) | 112 (8.0%) | 54 (9.0%) | 58 (7.4%) | 0.396 |
| Boat Accident | 5 (0.4%) | 3 (0.5%) | 2 (0.3%) | 1.00 |
| Buggy Accident | 28 (2.0%) | 15 (2.6%) | 13 (1.6%) | 0.563 |
| Blast Injury | 2 (0.1%) | 0 (0.0%) | 2 (0.3%) | 1.00 |
| Head Trauma | 82 (6.0%) | 39 (6.7%) | 43 (5.4%) | 0.264 |
| Loss of Consciousness<br>(Consequence of an injury) | 3 (0.2%) | 1 (0.2%) | 2 (0.3%) | 1.00 |
| Not Mentioned | 12 (0.9%) | 7 (1.2%) | 5 (0.6%) | 0.705 |
| Poly Trauma | 4 (0.3%) | 2 (0.3%) | 2 (0.3%) | 1.00 |
| Assault | 24 (1.8%) | 9 (1.6%) | 15 (1.9%) | 0.965 |
| Self-Inflicted Injury<br>(Gunshot) | 2 (0.15%) | 1 (0.2%) | 1 (0.1%) | 1.00 |

<sup>‡</sup> Chi-Square Test; WBCT, whole body computed tomography; RTA, road traffic accident; FFH, fall from height

**Fig. 1.**
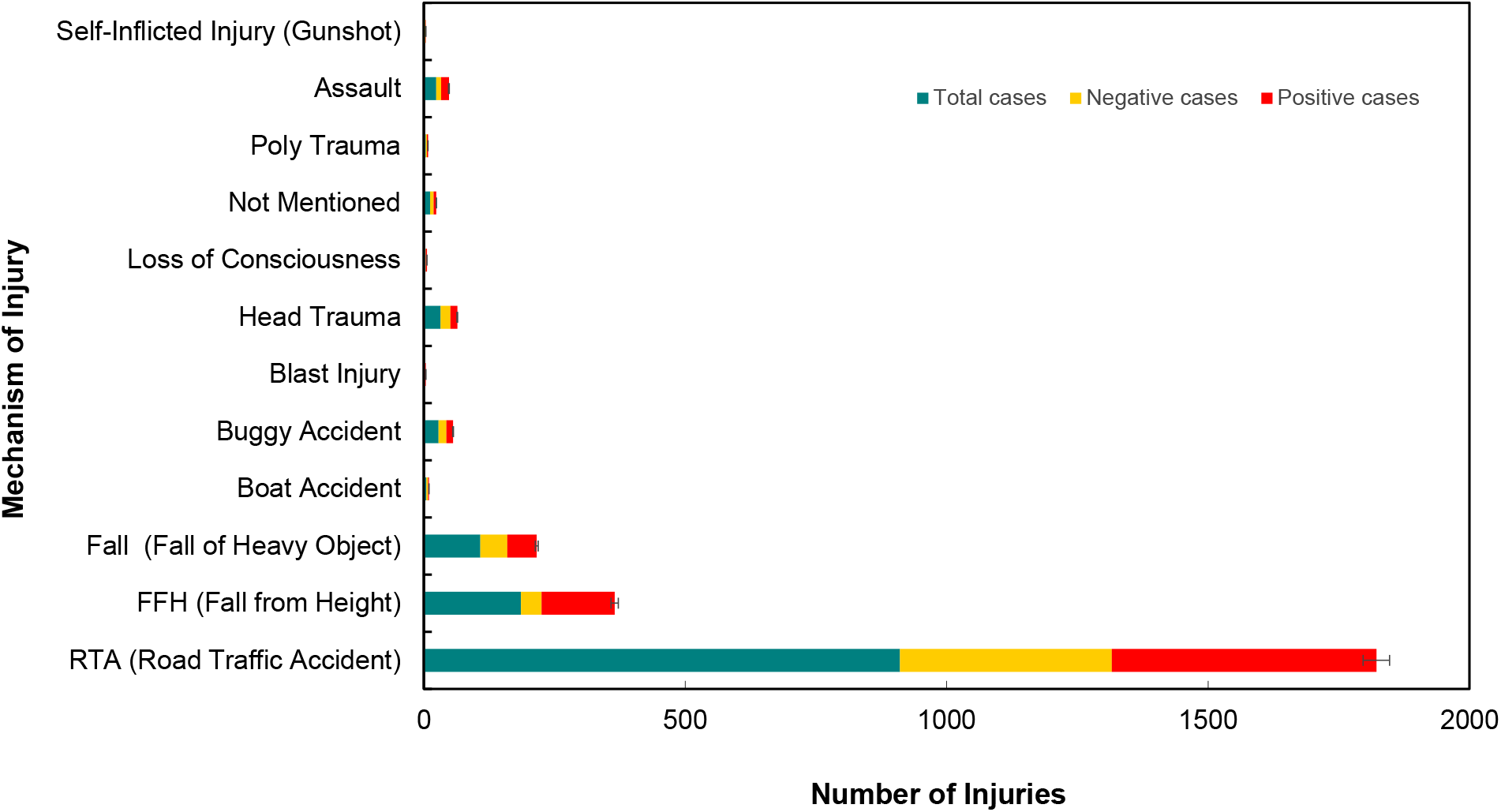
Subset trauma Classifications and the rate of indications.

**Fig. 2.**
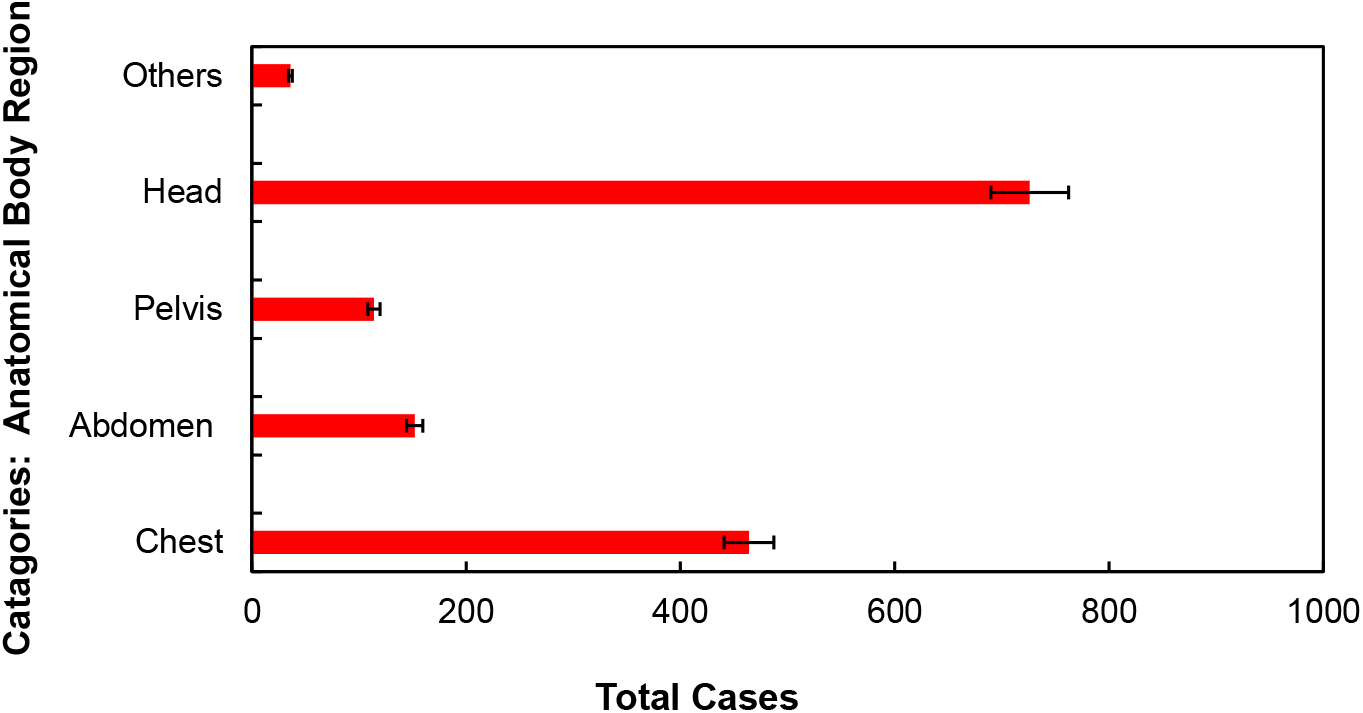
Anatomical body regions affected according to WBCT.

Description of WBCT scan finding and classification is given in Table 3, where the most common (top 5) finding injuries across all the collaborative radiology departments were reported to be: head hemorrhage/edema/hematoma 293 (21.4%), fracture spine 242 (17.7%), fracture ribs/clavicle/scapula 191 (14.0%), fracture skull/face 173 (12.7%) and lung contusions 162 (11.9%). For the positive case category, the mean values for CTDI (Computed Tomography Dose Index) volume, scan length, and DLP (Dose Length Product) were recorded as 89.01 ± 59.80 mGy, 43.53 ± 30.13 cm, and 2410.52 ± 1112.07 mGy x cm, respectively. The effective dose was calculated, yielding a mean ED of 22.41 ± 10.35 mSv. In comparison, for patients with negative CT findings, the mean ED was 19.98 ± 10.26 mSv, which significantly overlapped the range of positive CT cases (22.41 ± 10.35 mSv), considering the standard deviations of ±10.26 and ±10.35 (Table 4). Box plot comparisons of WBCT ED data across departments, showing overall consistency in dose distribution, with one department exhibiting greater variability in individual patient data, is illustrated in Figure 3. The limited availability of data may have influenced the variability.

**Table 3.** Description of WBCT Scan findings and classifications.

| **WBCT finding** |  | **(n=1367) - N (%)** |  |
| --- | --- | --- | --- |
| **Chest** | Lung contusions | 162 | (11.9%) |
|  | Fracture ribs/clavicle/scapula | 191 | (14.0%) |
|  | Pneumothorax | 85 | (6.2%) |
|  | Hemothorax | 19 | (1.4%) |
|  | Mediastinal hematoma | 7 | (0.5%) |
| **Abdomen** | Hepatic injury | 52 | (3.8%) |
|  | Splenic injury | 36 | (2.6%) |
|  | Renal injury | 12 | (0.9%) |
|  | Pancreatic injury | 2 | (0.1%) |
|  | Hemoperitoneum | 14 | (1.0%) |
|  | Mesenteric injury | 12 | (0.9%) |
|  | Adrenal injury | 5 | (0.4%) |
|  | Retroperitoneal hematoma | 16 | (1.2%) |
|  | Intestinal injury | 3 | (0.2%) |
| **Pelvis** | Pelvic fracture and/ hematoma | 112 | (8.2%) |
|  | Urinary bladder rupture | 2 | (0.1%) |
| **Head** | Fracture spine | 242 | (17.7%) |
|  | Brain hemorrhage/edema/hematoma | 293 | (21.4%) |
|  | Fracture skull/face | 173 | (12.7%) |
|  | Scalp hematoma | 18 | (1.3%) |
| **Others** | Long bone fracture | 29 | (2.1%) |
|  | Muscular injury | 2 | (0.1%) |
|  | Vascular injury | 5 | (0.4%) |

**Table 4.** Radiation characteristics and effective dose (ED) in trauma patients: Analysis across participating radiology departments.

|  | **Study cases**<br>**(n=1367)** | **Negative scan**<br>**(n=578)** | **Positive scan**<br>**(n=789)** | **p-value<sup>‡</sup>** |
| --- | --- | --- | --- | --- |
| **CTDIvol, (mGy)** | 84.33 ±55.80<br>(8.0 - 596.0) | 77.90± 49.11<br>(8.0-374.0) | 89.01± 59.80<br>(9.0-596.0) | 0.004 |
| **Scan Length (cm)** | 44.33 ±31.01<br>(5.0 - 153.0) | 45.45± 32.17<br>(7.0-150.0) | 43.53± 30.13<br>(5.0-153.0) | 0.520 |
| **DLP (mGy x cm)** | 2299.87 ±1115.28<br>(275.0 - 8671.0) | 2148.14± 1102.74<br>(275.0-6325.0) | 2410.52± 1112.07<br>(354.0-8671.0) | <0.001 |
| **ED- Effective**<br>**Dose (mSv)** | 21.38 ±10.37<br>(2.56- 80.64) | 19.98± 10.26<br>(2.56-58.82) | 22.41± 10.35<br>(3.29-80.64) | <0.001 |

<sup>‡</sup> Mann- Whitney U test; ED, effective dose; CTDIvol, computed tomography dose index-volume; DLP, dose length product

**Fig. 3.**
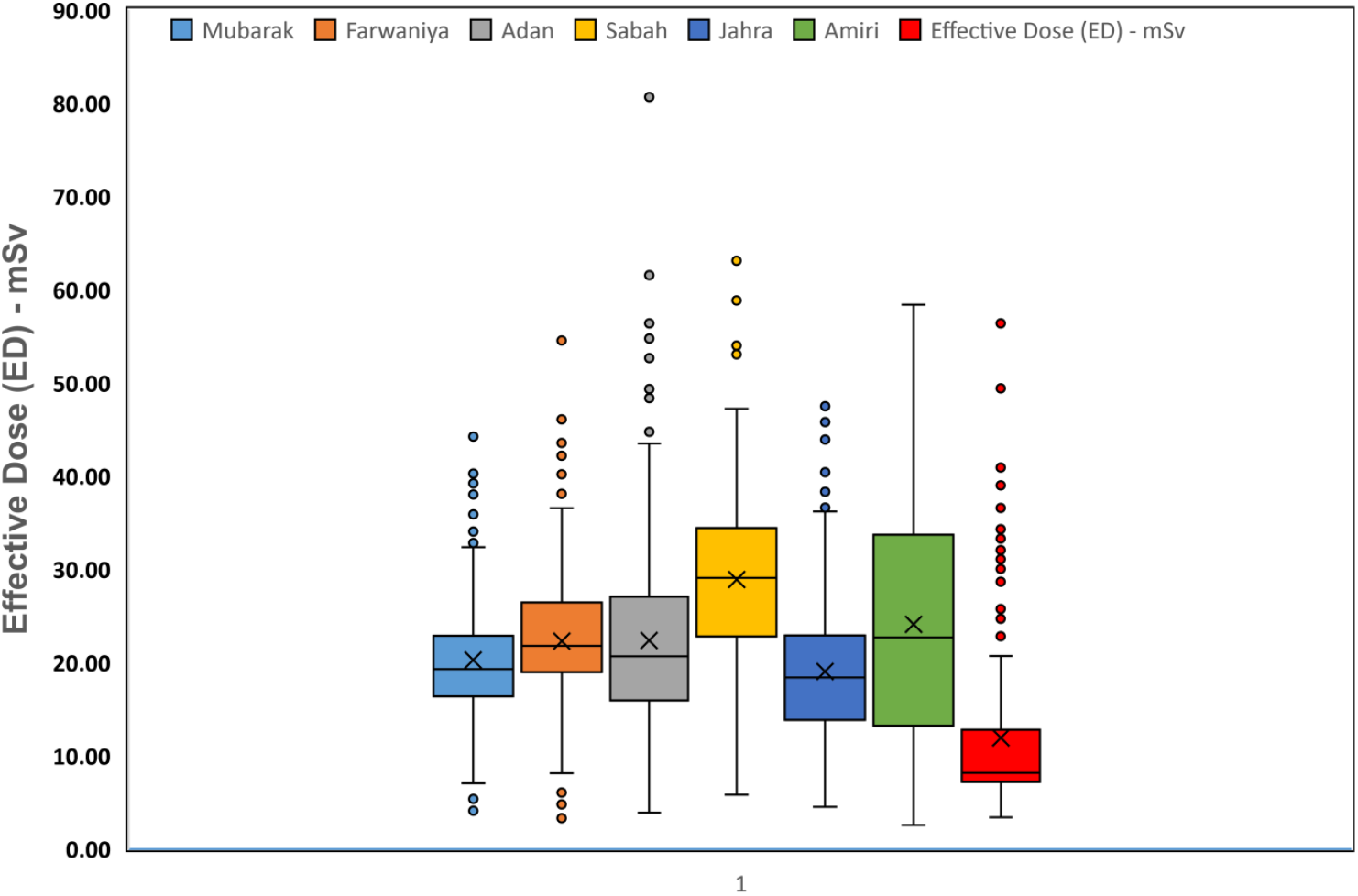
Box plot comparing WBCT effective dose (ED) across each radiology department, showing the overall pattern of dose distribution.

## 4. Discussion

WBCT plays a crucial role in detecting injuries and assessing their severity for treatment planning. According to a study by Kalra et al. [23], up to 22% of injuries may be missed without WBCT; however, these findings are often derived from studies conducted in major trauma centers in the United States, which typically deal with a combination of blunt and penetrating trauma and a higher volume of severe cases [24]. Furthermore, there is no conclusive evidence that WBCT reduces trauma mortality [25]. To the best of our knowledge, this is the first large-scale multicenter study in the State of Kuwait to evaluate the use of WBCT in trauma patients within emergency departments.

The occurrence of incidental findings, or “incidentalomas,” on CT scans has been reported in multiple studies, with some suggesting an incidence rate exceeding 50% [26,27]. A recent study from the New York trauma registry [28] found that 40% of trauma CTs had incidental findings. Of these, 63% were classified as Class 2 findings, which did not necessitate immediate attention but were not considered normal variations, thus requiring further investigation. Such incidental findings can be valuable for patients by enabling the early detection of significant underlying conditions. However, they may also lead to increased anxiety and healthcare costs due to additional investigations into abnormalities that may ultimately have no impact on the patient’s health [29-30]. Fear of missing significant pathology resulting in serious complications or outcomes and the worry of medicolegal ramifications are legitimate concerns [31].

A study by Caputo et al. [32] comparing the severity of injury, found that those undergoing WBCT had significantly higher Injury Severity Score (ISS) greater than 15 compared with those receiving selective scanning (29.72 vs. 26.46, p < 0.001, n = 23172). This might be misinterpreted as patients with more significant injuries being more likely to receive a WBCT scan, suggesting potential selection bias. However, both groups had ISS greater than 15, indicating that they were comparably severely injured. Meta-analyses of retrospective observational studies have reported that WBCT improves time to diagnosis and treatment, and reduces mortality in trauma patients, although these studies were heterogeneous and carried a high risk of bias [33,34]. Whole-body computed tomography for trauma has demonstrated a sensitivity of 95% and a specificity of 56% for detecting significant injuries [35].

The Randomized Study of Early Assessment by CT Scanning (REACT-2) is the only multicenter randomized controlled trial examining the use of WBCT in trauma. The trial included 1,403 severe trauma patients, randomized to either immediate WBCT or initial assessment with history, examination, chest X-ray (CXR), and FAST, followed by selective CT based on identified injuries. The study found no significant difference in mortality in 24 hours or 30 days. Subgroup analysis of polytrauma patients also showed no difference in mortality. However, REACT-2 demonstrated a reduction in time to diagnosis or treatment by 7 minutes overall, and 11 minutes in polytrauma patients. Despite some criticism, REACT-2 remains the highest quality evidence (level 2) for WBCT use in trauma [37]. Davies et al. [36] proposed a clinical scoring system considering multiple factors such as trauma to various body regions, Glasgow Coma Scale, hemodynamic and respiratory abnormalities, and injury mechanism. Several studies suggest using clinical prediction rules to safely omit unnecessary WBCTs, reducing radiation exposure and costs [29, 38-39]. While WBCT is beneficial for severe polytrauma, high rates of WBCT in trauma patients with no acute findings raise concerns about overuse, unnecessary radiation exposure, and incidental findings requiring follow-up [40].

In recent retrospective studies of hemodynamically stable patients after high-risk motor vehicle collision, low rates of injury requiring treatment or follow-up were found in body regions without clinically suspected injury, suggesting that CT should perhaps be selected based on physical findings [41]. An example of this can be found in the study by Huber-Wagner et al. [38], where 3,924 trauma patients underwent computed tomography as part of their initial evaluation. Of these, 2,430 patients received organ-selective CT, while 1,494 underwent whole-body CT. The study found that the predicted mortality rate was significantly higher in the whole-body CT group (23.2% vs. 17.3%; p < 0.001) compared to the organ-selective CT group (17.1% vs. 17.5%; p = 0.66) [35]. These findings were further explored in a follow-up study conducted by the same authors in 2013, which included a total of 16,719 patients (9,233 in the whole-body CT group and 7,486 in the organ-selective CT group). The study revealed that the absolute mortality rate in the whole-body CT group was significantly lower than in the organ-selective CT group (17.4% vs. 21.4%; p < 0.001) [38]. Several meta-analyses have supported these results, reinforcing the idea that whole-body CT is a safe, effective, and efficient tool for surgical triage [32,34,35,42]. The inclusion of a whole-body CT protocol in the management of severely injured trauma patients has been shown to reduce time to a definitive diagnosis, improve the accuracy of patient selection for operative versus non-operative management, and lower overall radiation exposure.

In our study, road traffic accidents (RTA) were the leading cause of injury among male patients (1074, 78.6%) and female patients (293, 21.4%) referred to ER departments. The overall rate of positive WBCT findings was 57.7%, with major cases at 38.8% and minor cases at 18.9%. While these findings provide valuable insights, they may lead to unnecessary follow-up investigations, increasing healthcare costs. A more selective approach would optimize resource use and allow for better allocation of funds to critical services. In countries like the UK, Germany, and Denmark, WBCT is applied more selectively, guided by clinical justification and reducing unnecessary radiation exposure [21].

In this study, we found that 42.3% of all WBCT findings were negative, which raising concerns about the unnecessary use of WBCT in radiology departments in Kuwait. This overuse can place significant strain on the 24/7 availability of skilled personnel and clinical services. In contrast, European countries such as the UK and Germany have reported lower percentages of negative findings (<20%) [43]. Additionally, we found that 73.9% of negative CT findings were associated with male patients, while 26.1% were associated with female patients. This discrepancy can be attributed to demographic factors, occupational risk exposure, and cultural norms. In Kuwait, males are more likely to engage in high-risk activities such as manual labor and driving, which results in a higher number of trauma cases requiring WBCT. Conversely, cultural norms may expose fewer females to such risks, thereby reducing their need for WBCT and contributing to the lower percentage of negative findings among them. This disparity reflects differences in trauma exposure demographics rather than overuse or inefficiency.

Although no large-scale epidemiologic studies of cancer risk have been reported in association with CT scans, experts state that up to 2% of cancers in the United States may be attributed to radiation exposure from CT scans. The stochastic or risk of chance mutations of carcinogenesis is suggested to be a linear relation between dose and biologic effect with no safe threshold [44]. In our study, the related ED among the participated radiology departments were in the range of 11.92 to 28.9 mSv that are influenced by including the type of CT scanner, acquisition parameters, and the protocol in use. Patients with negative CT findings, with a mean ED of 19.98 ± 10.26, are exposed to radiation levels within the same range as those with positive CT findings (mean ED of 22.41 ± 10.35). The overlapping standard deviations (±10.26 and ±10.35) highlight that negative findings result in radiation exposure similar to positive findings, raising concerns about unnecessary radiation exposure in these patients.

Two systematic reviews and analyses by Healy et al. [45] and Sierink et al. [46] found no difference in radiation mortality when comparing WBCT versus selective scanning of n = 8,180 and n = 5,470 trauma cases. To avoid unnecessary exposure and minimize the effect, adherence to the national diagnostic reference level (NDRL) which is set for CT diagnostic imaging based on the current facilities and practice in State of Kuwaitis recommended. In view of further optimisation and improving dose management, use of “Dose Watch” software solution that is a comprehensive, proactive radiation management program is recommended which can report the radiation dose delivered to patients during an examination and presents this data in an organized manner.

Kuwait’s national healthcare goals, as outlined in Vision 2035 [47], focus on improving healthcare services and maximizing resource efficiency. The unnecessary use of WBCT conflicts with these objectives by increasing costs and diverting resources from other critical healthcare initiatives. This highlights the need for stricter adherence to clinical guidelines to ensure WBCT is used only when it offers clear benefit, aligning trauma care with Kuwait’s broader healthcare vision. Additionally, data from WBCT can help shape public health strategies, particularly in addressing high-risk factors like road traffic accidents, a major trauma cause in Kuwait.

## 5. Limitations

Some limitations of our study include its retrospective design and the lack of consideration for injury severity. However, the impact of injury severity on the findings is likely minimal, as the two primary parameters—CT scan duration and image quality—are generally not affected by the patient’s condition.

## 6. Conclusions

In this study, 57.7% of WBCT findings were positive, with major cases accounting for 38.8% and minor cases for 18.9%. While these findings provide valuable insights, they may lead to unnecessary follow-up investigations, placing strain on healthcare resources. The rate of negative findings (42.3%) was significantly higher than the 20.4% average reported in similar centers in the UK and Germany. Among negative findings, 73.9% were male, and 26.1% were female, reflecting demographic factors, occupational exposure, and cultural norms. Road traffic accidents (RTA) were the leading cause of injury. Although mortality rates were not addressed, appropriate selection criteria are crucial to minimize unnecessary scans. The overlapping mean ED for both positive and negative CT scans raise concerns about the necessity of WBCT, particularly for negative findings. Adherence to the national diagnostic reference level (NDRL) for CT imaging in State of Kuwaitis recommended. Additionally, WBCT data can play a pivotal role in shaping public health strategies, particularly addressing road traffic accidents, a major trauma cause in Kuwait.

## Data Availability

All data produced in the present work are contained in the manuscript

## Authorship contributions

**Latifa Al-Kandari**: Conceptualization, Methodology, overall setup and joint funding acquisition. **Michael Masoomi**: Data curation, editorial, funding acquisition, project administration, Writing-Original draft, writing review and editing communication.

## Acknowledgements

The authors would like to acknowledge Hany Elrahman from the Medical Imaging Department at ADAN Hospital for his invaluable contribution in providing technical, computational services, and expertise. Furthermore, the authors express their appreciation to the following radiology departments in the State of State of Kuwait for their participation in providing trauma patient data (listed alphabetically): Al Adan Hospital – Ahmadi Health District; Farwaniya Hospital – Farwaniya Health District; Al-Jahra Hospital – Al Jahra Health District; Sabah Hospital – Al Sabah Specialized MED Center; Mubarak Al Kabeer Hospital – Hawally Health District; Amiri Hospital – Capital District; and Jaber Hospital – Capital District.

